# Long-term outcomes of radiofrequency catheter ablation for patients with persistent atrial fibrillation lasting ≥ 3 years

**DOI:** 10.1101/2024.04.11.24305700

**Authors:** Zikan Zhong, Jie An, Jiaqi Shen, Qitong Zhang, Xiaoyu Wu, Longzhe Gao, Yong Wei, Songwen Chen, Xiaofeng Lu, Juan Xu, Yu Ding, Cheng Cheng, Lidong Cai, Min Xu, Shaowen Liu, Genqing Zhou

## Abstract

**Background:** Radiofrequency catheter ablation (RFCA) for long-standing persistent atrial fibrillation (LSP-AF) remains challenging, especially in patients with very long atrial fibrillation (AF) duration.

**Objective:** To evaluate the long-term outcomes of RFCA in patients with LSP-AF lasting ≥ 3 years, and to identify predictors for AF recurrence.

**Methods:** This retrospective study included 151 patients with LSP-AF undergoing first-time RFCA. Procedure was performed with wide antral pulmonary vein isolation (PVI) based individualized ablation strategy, guided by ablation index (AI). Patients were followed up for ≥ 18 months, and recurrence predictors were determined.

**Results:** Enrolled patients (mean persistent AF duration: 7.6 ± 5.2 years) had a mean age of 65.3 ± 9.2 years and the median left atrial diameter (LAD) was 45.0 (42.0-49.0) mm. PVI was achieved in all, followed by modified left posterior wall isolation (PWI) in 147 patients. Additional ablation after PWI was performed in 88 patients. During the 18-month follow-up, the overall success rate was 74.2%. Multivariate analysis identified AF duration (HR 1.078; 95% CI 1.020-1.139; P = 0.007), LAD (HR 1.069; 95% CI 1.010-1.132; P = 0.022), and pre-procedure CRP (HR 1.063; 95% CI 1.010-1.117; P = 0.018) as independent predictors of AF recurrence. Among patients with PVI and PWI, those without empirical additional ablation (EAA) had a lower but not statistically significant recurrence rate (18.6%) than those with EAA (31.8%, P = 0.076).

**Conclusion:** Among LSP-AF patients, the long-term efficacy of AI-guided RFCA is acceptable, especially in selected patients. However, EAA after PVI and PWI may be unhelpful.

## Introduction

Atrial fibrillation (AF) is a prevalent, sustained cardiac arrhythmia that significantly impacts patients’ lives and elevates risks of mortality, stroke, and heart failure (HF) [1]. With advancing age, the incidence of AF exhibits an upward trend [2]. Radiofrequency catheter ablation (RFCA) has emerged as a primary treatment for AF rhythm control, aiming to reduce AF episodes, enhance quality of life, and slow the progression from paroxysmal to persistent AF [3, 4]. While studies support RFCA’s effectiveness over antiarrhythmic drugs [5], it’s increasingly apparent that for long-standing persistent AF (LSP-AF), pulmonary vein (PV) isolation alone may be inadequate. RFCA for LSP-AF patients is challenging, with less convincing outcomes [6]. The optimal strategy for LSP-AF remains elusive, with various additional ablation approaches attempted [7]. Ablation Index (AI) is gradually entering clinical practice to guide RFCA, yet its application in effectively guiding ablation for LSP-AF patients with extended AF duration or compromised left atrial substrate is limited [8]. Identifying predictors for AF recurrence and implementing preventive interventions is crucial. Established risk factors for AF recurrence include age, comorbidities, AF duration, left atrial (LA) size, ablation strategy, and et al [9–11]. However, their predictive performance in LSP-AF warrants further investigation. This study aims to evaluate long-term outcomes of AI-guided RFCA in patients with persistent AF lasting ≥ 3 years and identify predictors for AF recurrence.

## Methods

### Study population

This study employed a retrospective cohort design. Patients who underwent RFCA for LSP-AF from April 2017 to December 2021 were enrolled. Inclusion criteria: (1) age > 18 years old; (2) confirmed diagnosis of LSP-AF for at least 3 years; (3) ablation procedure was guided by AI with contact force-sensing catheters. Exclusion criteria included: (1) patients with a history of valvular heart disease (e.g., valve replacement, moderate or severe mitral stenosis); (2) previous AF procedure or surgery (e.g., ablation procedures, Maze surgery). All patients provided informed consent for RFCA. Ethical approval was granted by the Ethics Committee of Shanghai general hospital.

### RFCA

A multidetector CT scanner was performed to provide a comprehensive evaluation of LA and PVs structural features. LA thrombus was excluded using transesophageal echocardiogram prior to the ablation procedure or intracardiac echocardiography (ICE) during procedure in all patients. The procedure was carried out under conscious sedation, with continuous infusions of midazolam and fentanyl. Factor Xa inhibitor drugs or dabigatran were hold for one dose pre-procedure, and oral warfarin anticoagulation was uninterrupted to maintain an international normalized ratio (INR) level at 2.0-3.0. After femoral vein catheterization, a steerable decapolar catheter (Triguy, ATP Medical, Shenzhen, China) was inserted into the coronary sinus (CS). Atrial septum puncture was performed twice, following a first dose of heparin administration (80-130 IU/kg), and maintaining the activated clotting time (ACT) between 300 to 350 s during the procedure. A force-sensing catheter (Thermo-Cool Smart-Touch or Thermo-Cool Smart-Touch SF, Biosense Webster, Inc.) and the PentaRay (Biosense Webster, Inc.) catheter were introduced into the LA, guiding by CARTO 3 mapping system (Biosense Webster, Inc.). Point-by-point ablation was performed by using a power-controlled mode, with power settings of 25-45 W, maintaining a flow rate of 17-30 mL/min for the Smart-Touch catheter and 8-15 mL/min for the Smart-Touch SF catheter. Wide antral circumferential pulmonary vein ablation was the cornerstone for all patients, employing automated tagging (VisiTag, Biosense Webster) with predefined settings, including a minimum catheter stability time of 3 s, maximum range of 2.5 mm, force-time integral of 25%, minimum force of 5 grams, and lesion tag size of 2-3 mm. Inter-lesion distance between two neighboring lesions was set at ≤ 5.0 mm, with AI values of PV antral ablation at 500-550 on the anterior and the roof wall, 380-400 on the posterior and the inferior wall. After PV isolation (PVI), modified left posterior wall isolation (PWI) was performed in all patients, except for those in which AF was terminated after PVI. Whether additional ablation after PVI and PWI should be performed was determined by the physician. The AI values of roof and mitral isthmus linear ablation was set at 500-550, however, the goal of AI for other additional ablation was determined by the physician. After the completion of the ablation strategy, attempts were made to cardiovert AF to sinus rhythm (SR) with direct electrical cardioversion (ECV). In case of encountering atrial tachycardia (AT), activation and entrainment mapping are performed to determine the mechanism of AT, followed by ablation at the arrhythmogenic foci or critical isthmus. Bidirectional conduction block between LA and PVs, and of linear lesions, was assessed and confirmed under SR. After that, intravenous infusion of isoprenaline and a programmed stimulation from CS were performed to induce coexisting tachycardias in all patients. The specific surgical procedure flowchart is shown in Figure 1. Details of procedure-related complications were collected, including death, atrioesophageal fistula, phrenic nerve palsy, severe PV stenosis, ischemic stroke pericardial tamponade, hematoma, pseudoaneurysm, and arteriovenous fistula at access sites.

**Figure 1.**
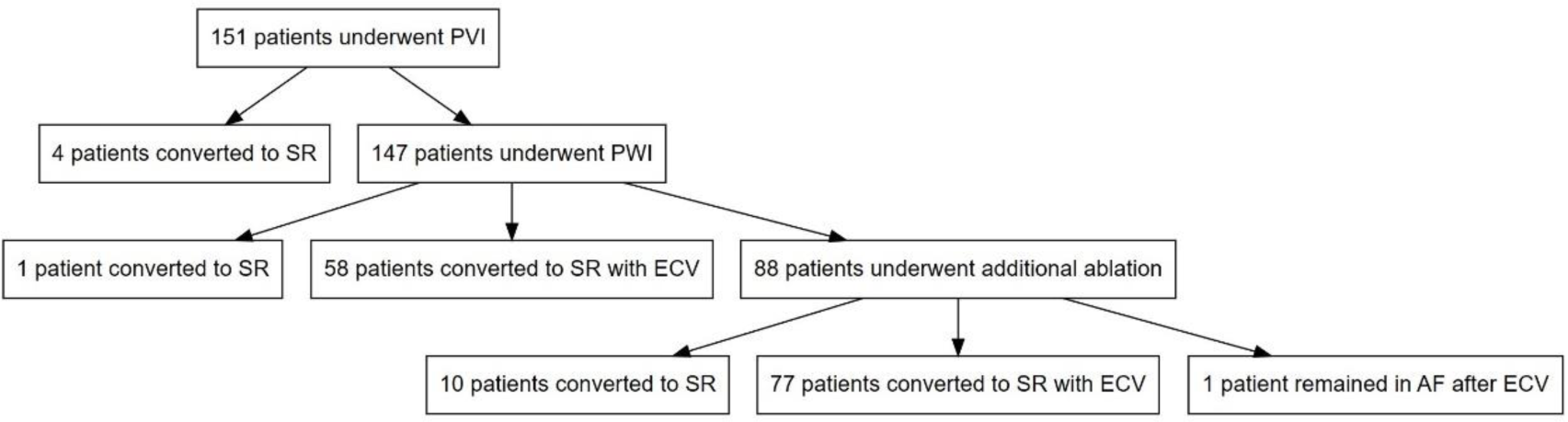
Schematic representation of the ablation procedure for PVI, PWI, and additional ablation steps in patients with long-standing persistent AF lasting ≥ 3 years. AF, atrial fibrillation; PVI, pulmonary vein isolation; PWI, posterior wall isolation.

### Follow-Up

Within the 3-month blanking period, patients received proton pump inhibitors for 1 month, and amiodarone or propafenone for 3 months if not contraindicated. The anticoagulants were administrated for at least 3 months, the decision on whether to continue using these medications was contingent on the CHA2DS2-VASc score. All patients were followed up at 3, 6, 12 months post-procedure, and every 6-month thereafter. The follow-up duration extended until July 2023. Additionally, all patients underwent 24-hour Holter monitoring at the respective follow-up time. Patients were advised to seek hospital admission for ECG upon the development of related symptoms. AF recurrence was defined as the presence of AF or AT episodes lasting ≥ 30 seconds after the 3-month blanking period. In cases where patients missed scheduled follow-up visits, they were contacted via telephone or other means to encourage timely adherence to the follow-up schedule. Safety endpoints during long-term follow-up were recorded, including cardiac death, stroke/transient cerebral ischemia (TIA), severe hemorrhage, and rehospitalization due to cardiac arrhythmia.

### Statistical Analysis

Continuous variables adhering to normal distribution were presented as mean ± standard deviation and analyzed using a two-independent-samples t-test. Non-normally distributed variables were expressed as median and interquartile range and analyzed via the Mann-Whitney test. Categorical variables were subjected to Pearson’s chi-squared test or Fisher’s exact test, and presented as numbers (percentages). Baseline clinical characteristic variables with p-values < 0.2 were included in a Cox regression analysis. Multivariate Cox regression was employed to identify independent predictors of AF recurrence post-RFCA. Receiver operating characteristic curves (ROC) were generated to establish cut-off values, area under the curve (AUC), sensitivity, and specificity for the variables. The risk of AF recurrence following RFCA was assessed using the Kaplan-Meier method, with statistical significance defined as P < 0.05. All statistical analyses were conducted using SPSS 25.0 (IBM Corp., Armonk, NY, USA) and GraphPad Prism (version 8.0).

## Results

### 3.1 Baseline Clinical Characteristics

This study included 151 patients with LSP-AF lasting over 3 years, of which 75.5% were male, with a mean age of 65.3 ± 9.2 years. The mean and median persistent AF duration were 7.6 ± 5.2 years and 5.0 (4.0 – 10.0) years, respectively; and the median left atrial diameter (LAD) was 45.0 (42.0 - 49.0) mm. A summary of baseline clinical characteristics is presented in Table 1. Among these patients, 96 (63.9%) had hypertension, 22 (14.6%) had coronary artery disease, 108 (71.5%) had HF, 42 (27.8%) had diabetes, and 15 (9.9%) had a history of stroke/ TIA. The mean CHA2DS2-VASc score was 2.9 ± 1.7, and the mean HAS-BLED score was 0.7 ± 0.6. A total of 128 patients (84.8%) were treated with novel oral anticoagulants, while the remaining patients received warfarin anticoagulation therapy.

**Table 1.**
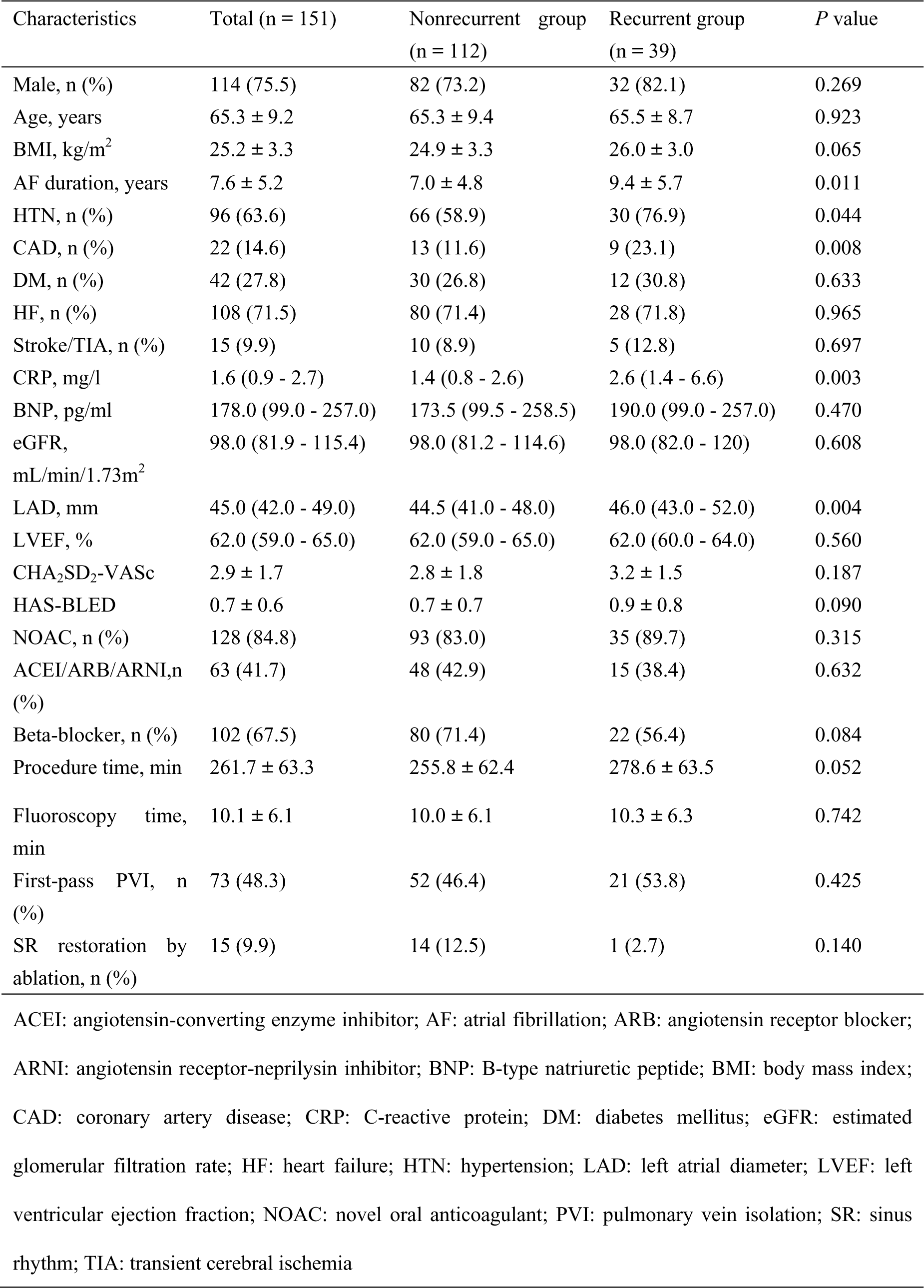
Characteristics of the study population and comparation between patients with or without recurrence.

### 3.2 Procedural details

All patients underwent successful PVI, and first-pass PVI was achieved in 73 (48.3%) patients. After PVI, AF spontaneously reverted to SR in 4 patients, the remaining 147 patients underwent modified PWI, with 59 receiving PWI only, and 88 underwent additional ablation after PWI. The mean procedure time was 261.7 ± 63.3 minutes, and the mean fluoroscopy time was 10.1 ± 6.1 minutes. Among the 59 patients who underwent PWI only (PWI group), one patient converted to SR during ablation of the LA roof line, while 58 patients remained in AF after PWI, and were successfully converted to SR after ECV. In the group of 88 patients who underwent empirical additional ablation (EAA) after PWI (PWI+EAA group), AF was terminated to SR in 4 patients, including 3 patients during ablation of the LA anterior wall line and 1 during superior vena cava isolation. Additionally, 6 patients experienced AF conversion to AT during further ablation: 3 during LA anterior wall line ablation, 2 during LA bottom line ablation, and 1 during cavotricuspid isthmus line ablation, the ATs were successfully terminated after further ablation. The remaining 78 patients who underwent PWI and EAA remained in AF and underwent ECV, with 77 patients successfully converting to SR. One patient failed to convert to SR after multiple attempts with ECV and the procedure was terminated. The overall incidence of periprocedural complications was 4.0% (6/151). There were 6 cases of puncture site bleeding. None of the patients experienced other complications.

### 3.3 Follow-up results

The overall success rate at 18-month follow-up was 74.2% (112/151), the Kaplan-Meier curve of arrhythmia-free survival in the study population is shown in Figure 2. When compared to patients without AF recurrence (nonrecurrent group, n = 129), patients with recurrence (recurrent group, n =46) had longer persistent AF duration (9.4 ± 5.7 vs. 7.0 ± 4.8 years, P = 0.011), higher CRP level [2.6 (1.4 – 6.6) vs. 1.4 (0.8 - 2.6) mg/L, P = 0.003], larger LAD [46.0 (43.0 - 52.0) vs. 44.5 (41.0 – 48.0) mm, P = 0.004], and longer procedure time (278.6 ± 63.5 vs. 255.8 ± 62.4 min, P = 0.052). Regarding comorbidities, the recurrent group exhibited a higher prevalence of hypertension (76.9% vs. 58.9%, P = 0.044) and coronary artery disease (CAD, 23.1% vs. 11.6%, P = 0.008) compared to the non-recurrent group. No significant differences in other baseline characteristics were observed between the two groups (Table 1). During the follow-up period, there was one case of acute stroke and one case of sudden cardiac death in the nonrecurrent group, as well as one case of acute upper gastrointestinal bleeding in the recurrent group. Additionally, in recurrent group, there was a higher proportion of patients who were readmitted during long-term follow-up, due to worsening symptoms related to AF events compared to the nonrecurrent group (35.9% vs. 7.1%, P < 0.001). In subgroup analysis of these 147 patients based on ablation strategy, the “PWI + EAA” group showed a higher 18-month recurrence rate compared to the “PWI” group, although it did not reach statistical significance [31.8% (28/88) vs. 18.6% (11/59); P = 0.076]. Furthermore, the “PWI + EAA” group had a longer procedure duration (278.6 ± 63.5 vs. 255.8 ± 62.4 min, P = 0.017). The Kaplan-Meier curve of arrhythmia-free survival according to different ablation strategy is shown in Figure 3. Additionally, there were no significant differences between the two groups in terms of baseline characteristics or the occurrence of long-term or short-term complications during post-procedure follow-up (Table 2).

**Figure 2.**
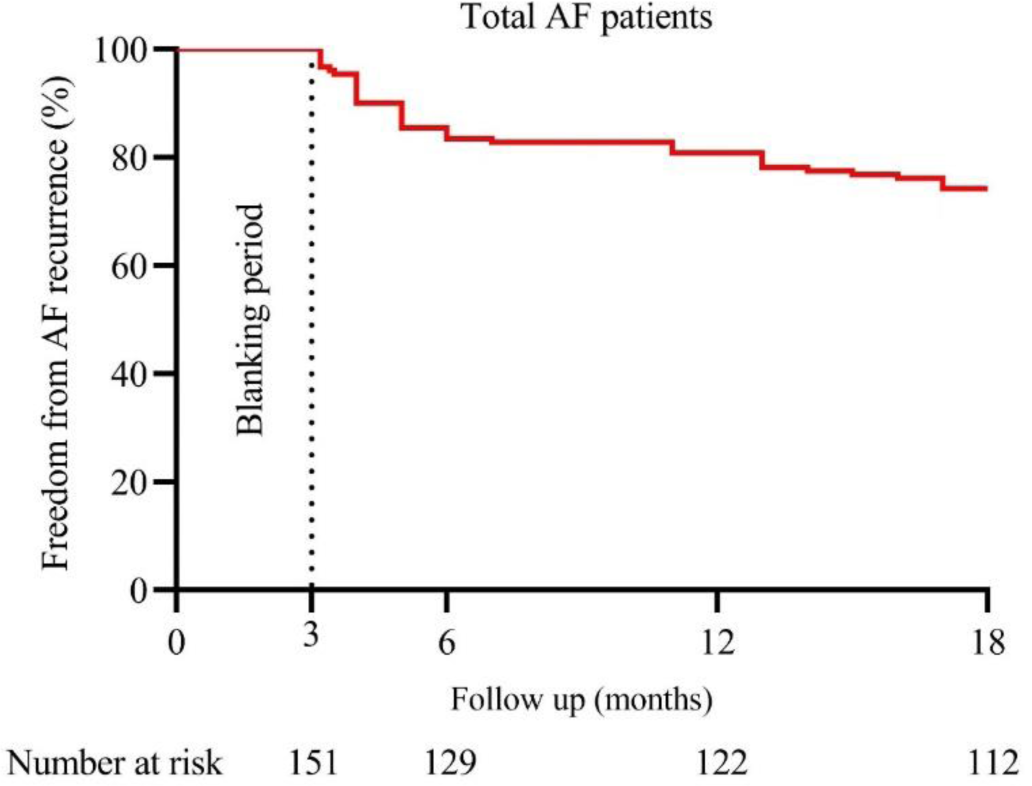
The Kaplan-Meier analysis illustrating the effectiveness of AI-guided radiofrequency catheter ablation procedure in patients with persistent AF lasting ≥ 3 years. At the 18-month follow-up, the overall success rate of catheter ablation was 74.2%. AF, atrial fibrillation; AI, ablation index.

**Figure 3.**
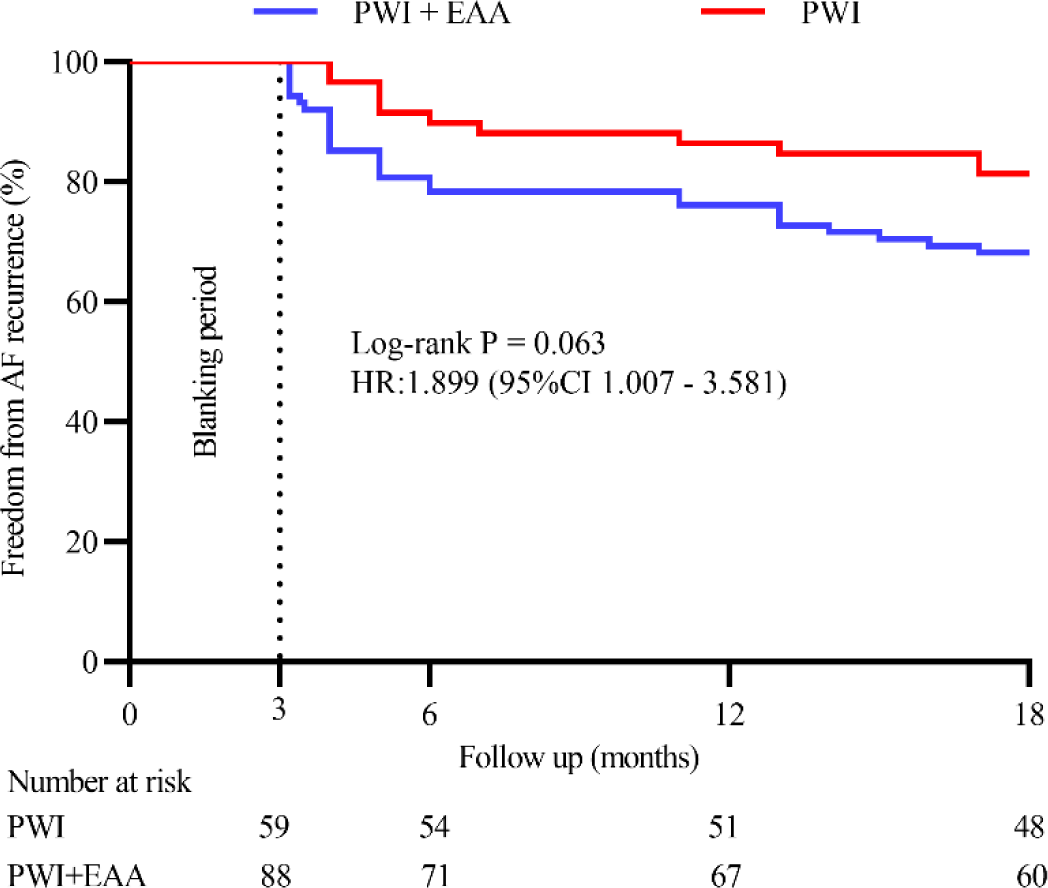
The Kaplan-Meier analysis of AF recurrence-free rates during follow-up based on ablation strategy. AF, atrial fibrillation.

**Table 2.**
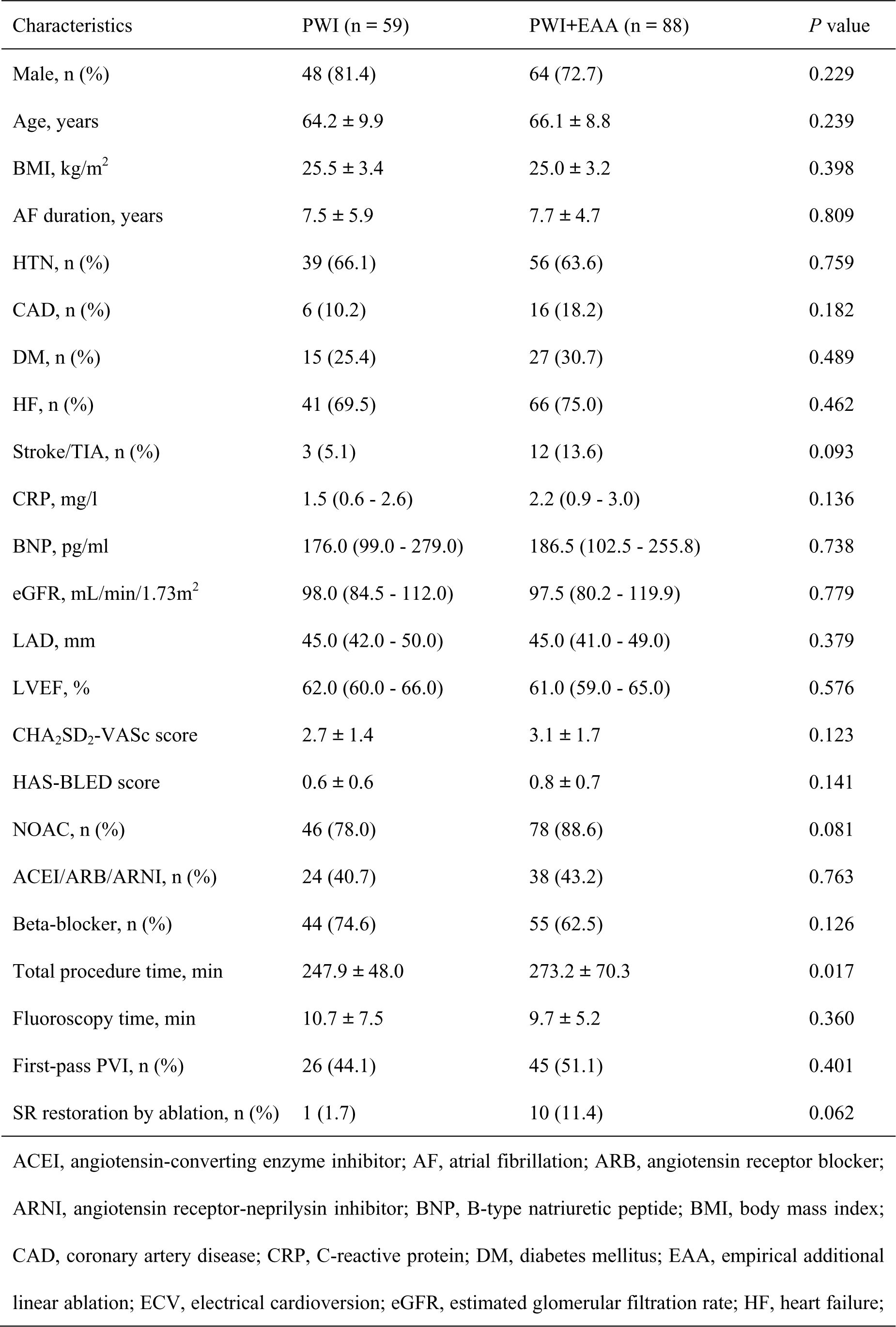
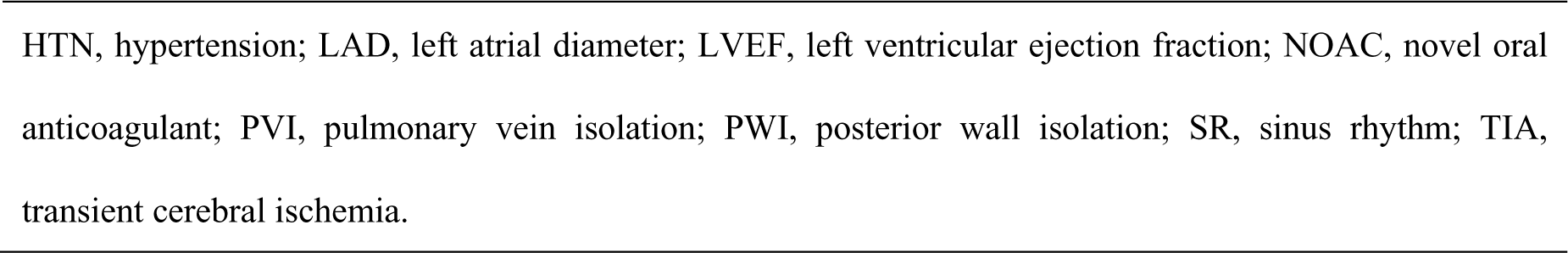
Comparison of “PWI” and “PWI+EAA” Groups in LSP-AF Patients.

### 3.4 Predictors of AF recurrence

Univariate Cox regression survival analysis revealed significant associations between multiple factors and AF recurrence, including AF duration (HR 1.068; 95% CI 1.018 - 1.121; P = 0.007), pre-procedure CRP level (HR 1.097; 95% CI 1.047-1.149; P < 0.001), LAD (HR 1.083; 95% CI 1.028-1.141; P = 0.003) and procedural time (HR 1.005; 95% CI, 1.001 - 1.010; P = 0.026). Further multivariate Cox regression analysis. confirmed that the AF duration (HR 1.078; 95% CI 1.020-1.139; P = 0.007), LAD (HR 1.069; 95% CI 1.010-1.132; P = 0.022) and pre-procedure CRP (HR 1.063; 95% CI 1.010-1.117; P = 0.018) were independently associated with AF recurrence (Table 3).

**Table 3.**
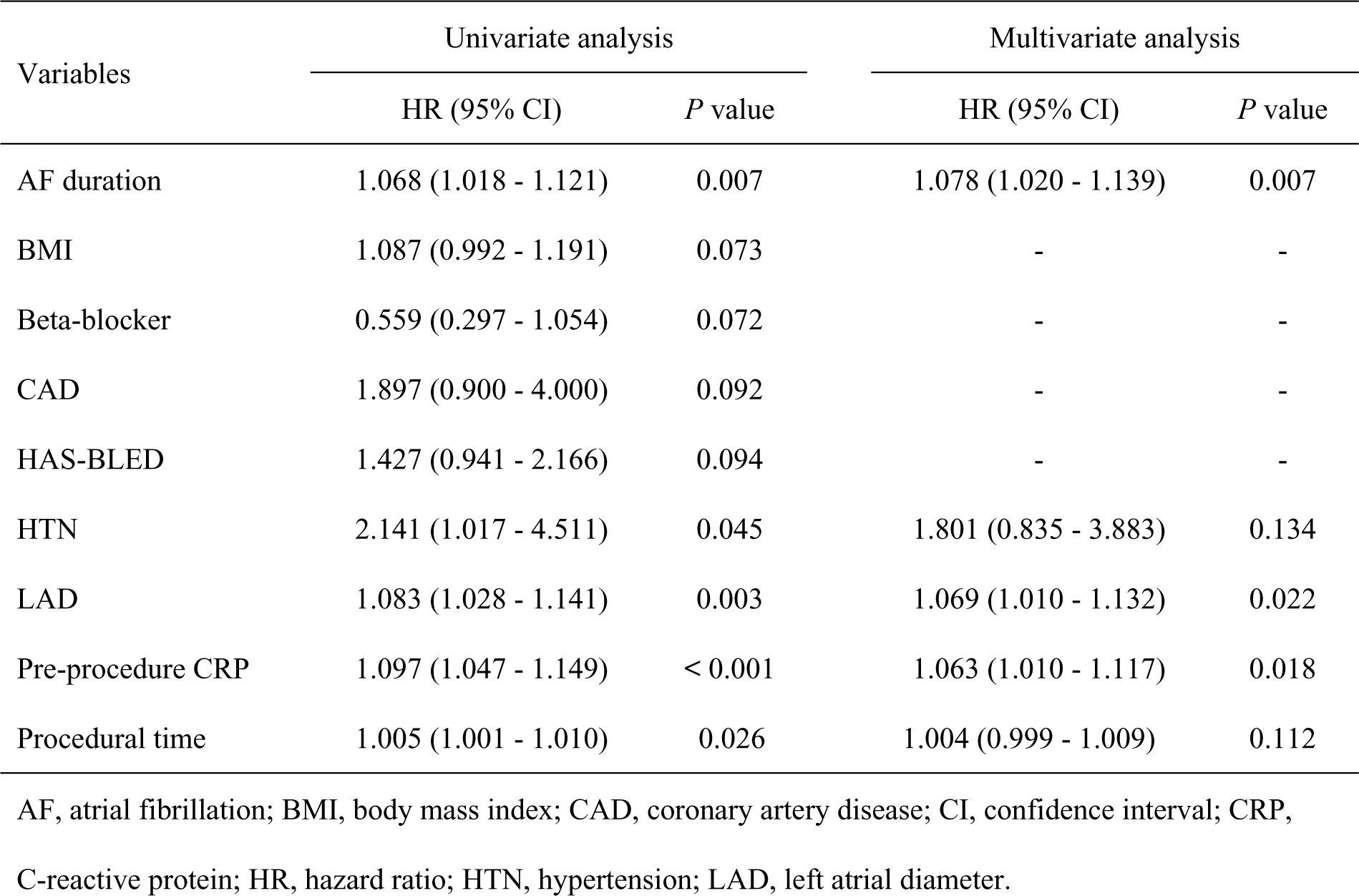
Univariate and multivariate Cox regression analysis of predictors for AF recurrence.

### 3.5 ROC Curve Analysis and Predictive Value of Predictors for AF Recurrence

Figure 3 illustrates the ROC curves for AF duration, LAD and pre-procedure CRP, with respective AUC values of 0.647 (95% CI 0.548 - 0.745; P = 0.007), 0.653 (95% CI 0.547 - 0.759; P = 0.005) and 0.658 (95% CI 0.552 - 0.765; P = 0.003). The cutoff value for AF duration was determined to be 4.5 years (sensitivity 84.0%, specificity 42.9%), while the cutoff value for LAD was 50.5 mm (sensitivity 35.9%, specificity 89.3%) and pre-procedure CRP was 2.0 mg/L (sensitivity 69.2%, specificity 62.5%). The AUC of combination of AF duration, LAD and pre-procedure CRP was 0.788 (95% CI 0.703 - 0.873; P < 0.001), with sensitivity at 79.5% and specificity at 71.4%.

Furthermore, Kaplan-Meier curves were employed to depict the time to AF recurrence for patients grouped based on different cutoff values of AF duration, pre-procedure CRP, and LAD in Figure 4. Notably, these subgroups included patients with CRP level above and below the 2.0 mg/L cutoff (P < 0.001), patients with AF duration exceeding and falling below 5 years (P = 0.002), and patients with LAD greater than and less than 50 mm (P = 0.002). (Figure 5)

**Figure 4.**
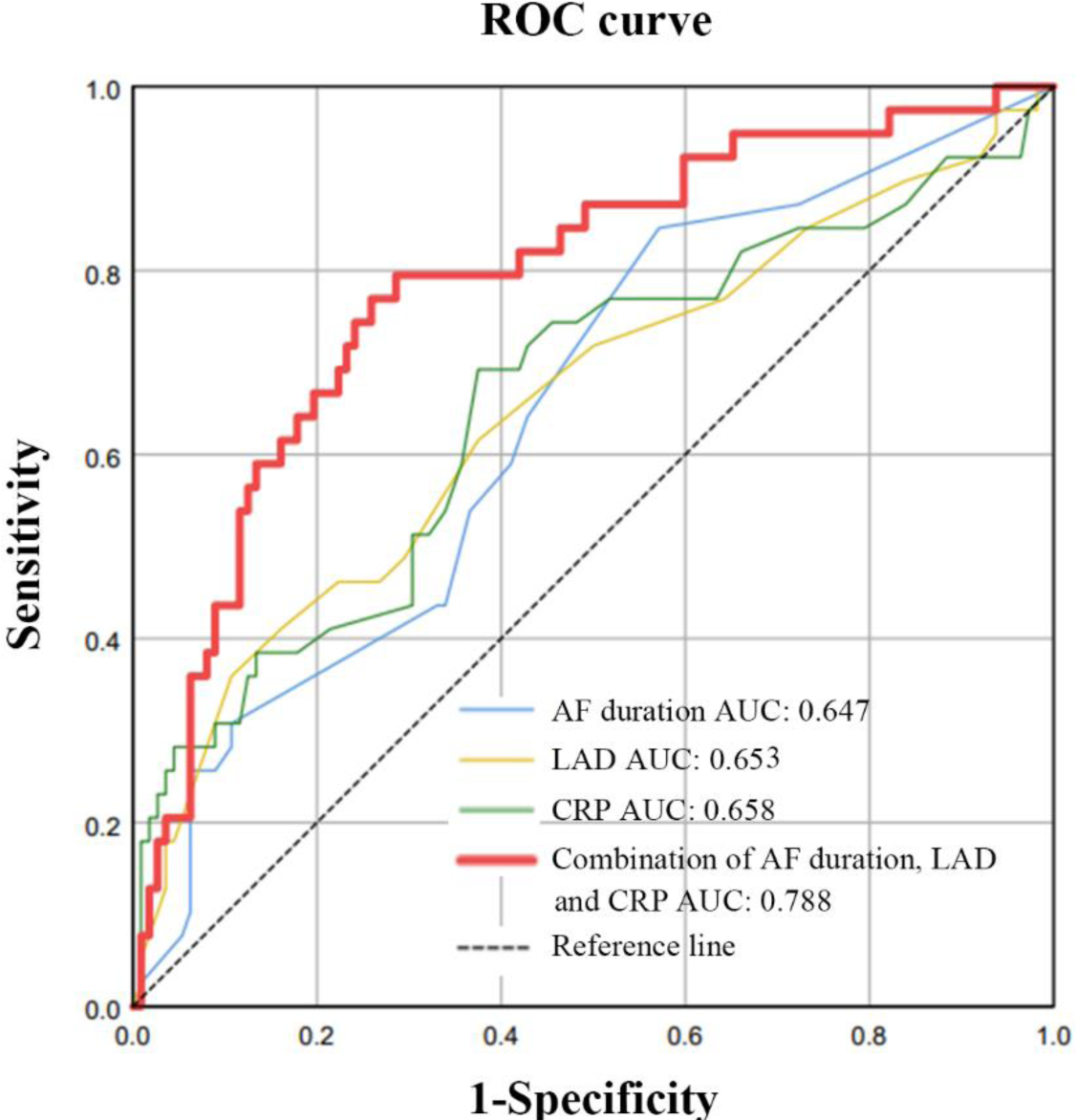
Determination of the predictive value of AF duration, CRP level, and LAD in AF Recurrence. The AUC = 0.647 for AF duration, P = 0.007; AUC = 0.658 for pre-procedure CRP, P = 0.003; AUC = 0.653 for LAD, P = 0.005; The AUC of combination of AF duration, LAD and pre-procedure CRP was 0.788, P < 0.001.

**Figure 5.**
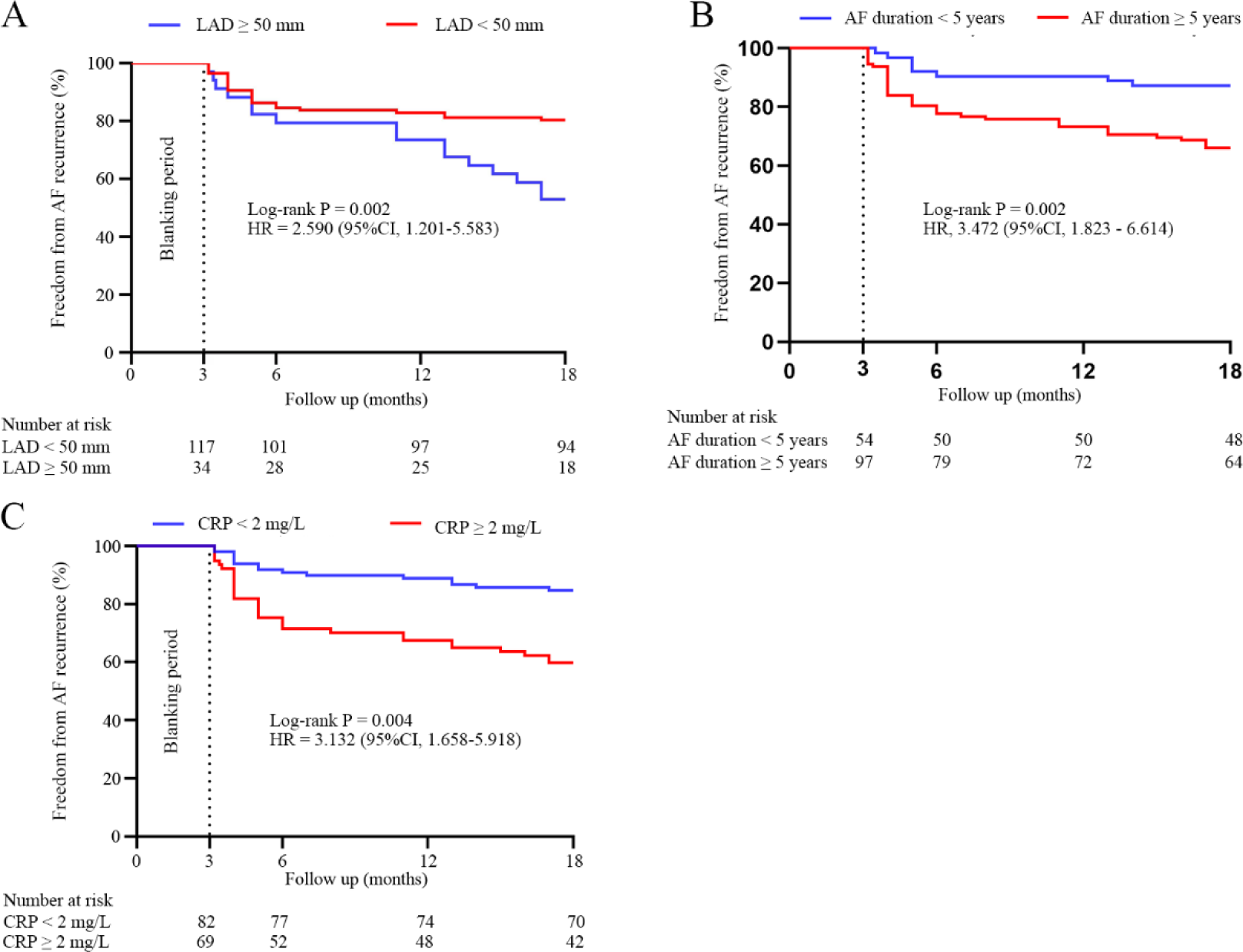
The Kaplan-Meier analysis of AF recurrence-free rates during follow-up based on specific cutoff values for AF duration, pre-procedure CRP level, and LAD (CRP ≥ 2.0 mg/L: P < 0.001; AF duration ≥ 5 years: P = 0.002; LAD ≥ 50 mm: P = 0.002). AF, atrial fibrillation; CRP, C-reactive protein; LAD, left atrial diameter;

## 4 Discussion

### 4.1 Main Findings

Focused on patients with LSP-AF lasting ≥ 3 years, our study investigated the long-term follow-up results and factors associated with AF recurrence following RFCA. The overall success rates at 18 months post-RFCA was 74.2% for LSP-AF patients. Notably, LAD, AF duration, and pre-procedure CRP level emerged as independent predictors of AF recurrence. Additionally, in patients with LSP-AF lasting ≥ 3 years, if AF persisted after PVI and PWI, EAA did not improve the long-term success rate of RFCA.

### 4.2 Value of LSP-AF Catheter Ablation

AF burden significantly impacts HF development, especially in LSP-AF, exacerbating cardiac workload. In our cohort of 151 LSP-AF patients, 108 (71.5%) presented with concomitant HF, highlighting the increased cardiac burden in this subgroup. AF and HF commonly coexist, mutually influencing each other [12]. The risk of HF in AF patients is 1 to 2 times higher than in those without AF, while HF patients have a 2-fold increased risk of developing AF [13]. In patients with HF with reduced ejection fraction, the prevalence of AF ranges from 36.7% to 44.9%, and in those with HF with preserved ejection fraction, the AF prevalence is between 40% and 50% [14]. Importantly, HF is a major cause of mortality in AF patients, as evidenced by a Chinese community study showing that HF is the leading cause of death in persistent AF patients [15]. A meta-analysis suggests that catheter ablation can improve the prognosis and quality of life of patients with AF combined with HF [4]. The CASTLE-HTx study revealed that catheter ablation for AF can reduce the composite endpoint in patients with end-stage HF, including total mortality, left ventricular assist device implantation, or the occurrence of HF worsening requiring urgent heart transplantation. This suggests that AF remains a significant risk factor for HF deterioration, even in end-stage HF patients, and can be corrected through catheter ablation, stabilizing their clinical condition [16]. Despite low success rates, catheter ablation still clinically benefits these patients by reducing AF burden and reversing AF types. Thus, RFCA remains beneficial in LSP-AF, offering potential to improve outcomes by alleviating AF burden.

### 4.3 Patient Selection and Predictors of Recurrence

AF is often likened to “cancer” in the realm of arrhythmias, with LSP-AF patients akin to late-stage metastatic cancer patients [17]. The concept of “AF begets AF” underscores the challenge of managing prolonged AF states [18]. Current research has demonstrated a significant correlation between pre-procedure AF duration and post-RFCA recurrence rates [19, 20]. One study has indicated that longer AF duration significantly increases the recurrence rate, especially among patients with persistent AF lasting over three years [10]. Our study further categorized patients based on AF duration, revealing a higher recurrence rate among those with AF duration exceeding 5 years. This result further underscores the association between AF duration and recurrence risk. Therefore, patients with exceptionally prolonged LSP-AF durations may face an elevated risk of AF recurrence.

The assessment of LA substrate has long been a critical criterion for evaluating the efficacy of RFCA procedures. Among the frequently employed metrics, LAD and left atrial volume (LAV) stand out as primary indicators. Notably, D’Ascenzo et al. reported that a LAD exceeding 50 mm was among the most potent predictors of AF ablation failure in the general population [21]. Additionally, numerous previous studies have consistently emphasized the significance of left atrial size as a significant predictive factor for AF recurrence post-ablation [22]. Our study confirms LAD’s critical role by categorizing patients based on LAD, showing higher AF recurrence rates in LAD ≥ 50 mm group. This underscores LAD’s significance in assessing AF recurrence risk in LSP-AF patients. It’s noteworthy that in Kohei Ukita’s study, there was a notably higher proportion of patients with persistent AF in the LAD ≥ 50mm group. Our study specifically targeted LSP-AF patients with a duration exceeding 3 years, enhancing its specificity [23]. In summary, our research highlights the importance of LAD as a critical predictor of late AF recurrence in LSP-AF patients. Chronic inflammation and immune infiltration are key contributors to AF onset and progression, influencing atrial tissue remodeling [24]. Serum biomarkers associated with inflammation, like CRP, hold potential in predicting AF recurrence post-ablation [25–27]. Our study, focusing on LSP-AF patients with > 3 years duration, found a notable association between elevated baseline CRP levels and AF recurrence. Specifically, a 1 mg/L increase in CRP corresponded to a 6.3% rise in recurrence rate, with the ≥ 2.0 mg/L group exhibiting a three-fold higher recurrence rate compared to < 2.0 mg/L. These findings underscore CRP’s predictive value in assessing AF recurrence risk following RFCA in this patient subset. In conclusion, our results suggest that assessing AF duration, pre-procedure LAD, and baseline CRP level aids in the preprocedural identification of patients at higher risk of recurrence, thereby improving electrophysiological assessment and ablation strategy decision-making.

### 4.4 Ablation Strategy for LSP-AF

In addition, a meta-analysis indicates a significant reduction in AF recurrence rates for patients with persistent AF through the use of PWI, especially in elderly individuals with larger left atria [28]. Furthermore, our center has proposed a modified posterior wall line to reduce esophageal injury while simultaneously isolating a more extensive area of the posterior wall and posterior-inferior atrial tissue region. By eliminating potential triggers and altering atrial electrophysiological remodeling, the efficacy of ablation is further enhanced [29]. However, the necessity of EAA following PWI remains a topic of debate in the management of persistent AF. In our study cohort, the “PWI + EAA” group exhibited a longer procedure duration. While the “PWI + EAA” group showed a slightly higher recurrence rate compared to the “PWI” group, this difference was not statistically significant (P = 0.076). Subgroup analysis suggests EAA may not add significant benefit in LSP-AF post-PWI. However, the observed difference in recurrence rates may be influenced by the extent of atrial fibrosis. While baseline blood test results and imaging findings were comparable between the two groups, there was a lack of quantifiable measurements of atrial substrate, such as the extent or distribution of low-voltage areas in the atria. This presents an avenue for future investigation. Additionally, achieving transmural lesions or bidirectional block in regions like the mitral isthmus during EAA execution can be challenging. Furthermore, for patients with LSP-AF and adverse atrial substrate conditions, mapping and successfully ablating triggering foci outside the PVs may be difficult during the procedure. These challenges could contribute to the limited efficacy of EAA in reducing recurrence rates in LSP-AF following PWI.

## 5. Limitations

Nevertheless, it is imperative to acknowledge the limitations inherent in our study. Our research bears the hallmarks of a retrospective observational design with a relatively modest sample size. To substantiate these findings, large-scale RCT remain imperative. Furthermore, our institutional reliance on transthoracic echocardiography for cardiac ultrasound parameters limited our assessment to the LA size. While informative, this metric may not offer the same comprehensive insight as a comprehensive evaluation of LAV. Additionally, the monitoring and assessment of patients were predominantly conducted through intermittent 24-hour holter monitoring at specific intervals. However, this intermittent monitoring approach has the potential to miss episodes of atrial arrhythmias occurring between monitoring sessions, leading to potential underestimation of recurrence rates, especially in patients with asymptomatic episodes.

## 6. Conclusions

Among patients with LSP-AF lasting ≥ 3 years, the long-term efficacy of AI-guided RFCA is acceptable, especially in selected patients. However, EAA may be unhelpful if AF persistent after PVI and PWI.

## Data Availability

The figure was created by SPSS 25.0 (IBM Corp., Armonk, NY, USA) and GraphPad Prism (version 8.0).

## Availability of Data and Material

The figure was created by SPSS 25.0 (IBM Corp., Armonk, NY, USA) and GraphPad Prism (version 8.0).

## Code Availability

Not applicable.

## Authors’ Contributions

Conceptualization, G.Z. and S.L.; investigation, L.G., X.W. and L.C.; writing—original draft preparation, Z.Z, J.S. and J.A.; writing—review and editing, J.X, X.L and G.Z.; supervision, Q.Z., Y.W., Y.D., X.L. C.C., M.X., and S.C.; All authors have read and agreed to the published version of the manuscript.

## Ethics approvals

Ethical approval was granted by the Ethics Committee of Shanghai general hospital.

## Consent to participate

Not applicable.

## Consent for publication

Not applicable.

## Supplementary data

**Supplementary Table 2.**
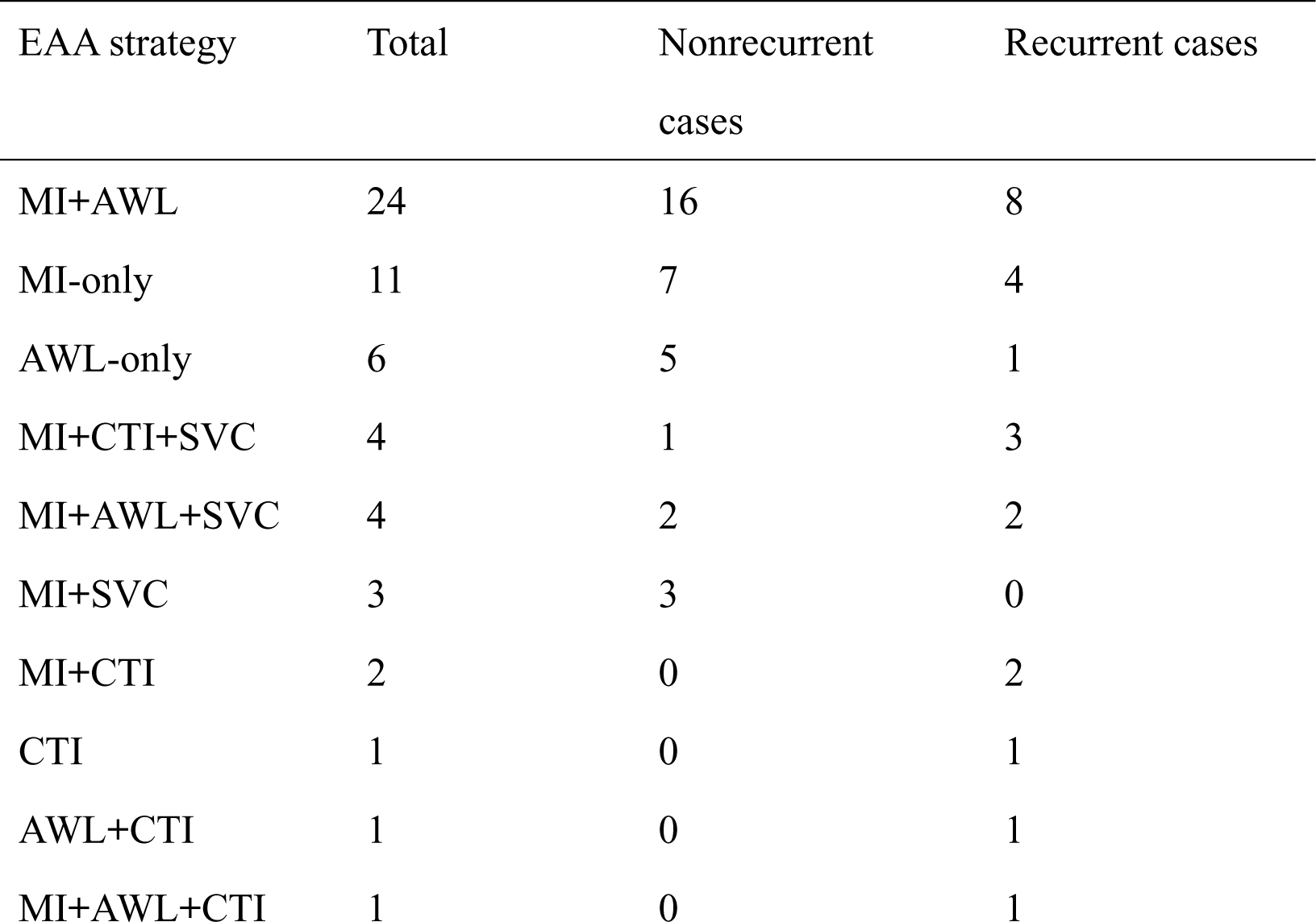
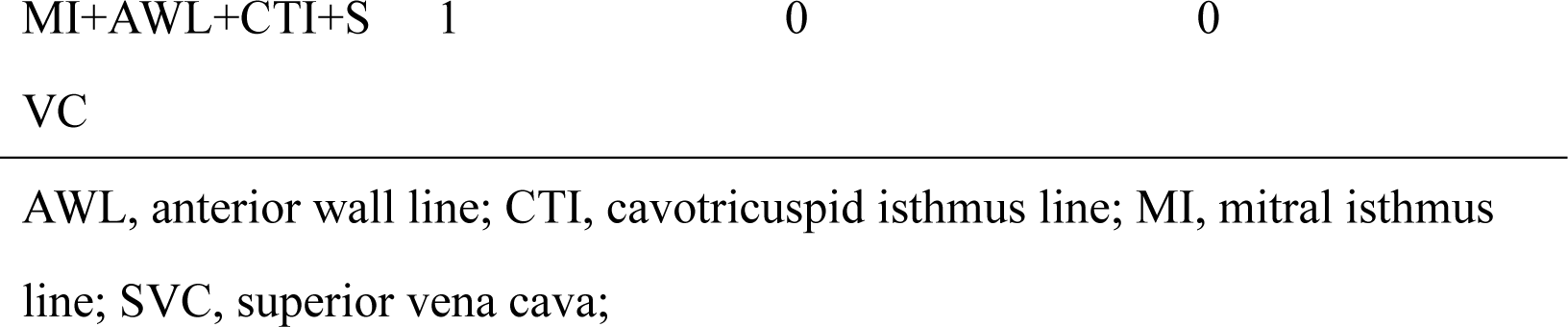
Strategies of EAA.

